# Can large language models extract travel history from clinical text? A multi-annotator benchmark study

**DOI:** 10.64898/2026.09.29.26363894

**Authors:** Praveenkumar Kanithi, Alyssa Pradhan, Haochen Cui, Kieran Johnstone, Augustine Luk, Robert Hammond, Colin Brown, Ann Sarah Walker, David Eyre

## Abstract

International travel can increase infection risk and spread antimicrobial resistance (AMR), yet clinical systems rarely capture travel history in a structured format. Extracting this information from free-text clinical documentation at scale could improve alerts for high-consequence infections and AMR surveillance. We developed an annotation framework covering key aspects of travel history, including destination, exposures, travel duration, and multiple temporal variables. The framework was used to annotate a corpus of 100 clinical case reports by five clinicians, yielding over 5,000 clinician annotations. Several LLMs were evaluated on the same extraction task, with their outputs compared against clinician consensus. Agreement among clinicians was high for straightforward variables such as presence of travel and destination, but dropped substantially for temporal variables such as time since return and symptom onset relative to return. LLMs showed a similar pattern. They performed well on high-level variables but struggled with the same temporal fields that challenged clinicians. Model errors were also concentrated in cases where clinicians disagreed. Across models, outputs were more sensitive to sampling temperature than to prompt wording. A smaller open-weight model performed similarly to substantially larger models on several variables but remained weaker on temporal fields. Overall, LLMs could extract some travel history variables, but reliable extraction of temporal information remained difficult.

## 1 Introduction

International travel has grown substantially over recent decades, with 1.5 billion international tourist arrivals recorded annually (UN Tourism, 2026; Théry, 2025). This mobility creates pathways for the importation and spread of infectious diseases and/or antimicrobial-resistance (AMR) determinants across geographic boundaries. Travel is an established risk factor for a range of infections including malaria, typhoid, dengue, and enteric pathogens, as well as for the acquisition and onward transmission of AMR organisms (Bokhary et al., 2021; Aggarwal et al., 2022). Travel history is therefore a central element of the clinical work-up for patients presenting with febrile illness or suspected infection, influencing both differential diagnosis and empirical treatment decisions.

Despite its clinical importance, travel history is rarely captured in structured form within electronic health records (EHRs) (Chapman et al., 2010; Peterson et al., 2021). When recorded, this information typically exists as free text embedded in clinical case notes, discharge summaries, or referral letters (Johnson et al., 2023b). The absence of structured travel data limits its utility for real-time alerting, e.g., prompting to test for malaria or identifying patients at risk of viral hemorrhagic fevers needing specific precautions, and also limits secondary uses including epidemiological surveillance, outbreak detection, and population-level analyses of travel-associated infection or AMR acquisition. Retrospective extraction of this information at scale requires either labour-intensive manual review (Nye et al., 2021), which is seldom feasible, or more tractably, automated approaches capable of processing unstructured clinical text (Lopez et al., 2025).

The specific elements of travel history that carry diagnostic and epidemiological relevance are well-defined. For any patient presenting with a travel-associated illness, clinicians seek to establish: whether recent travel occurred; the destination(s) visited; the duration of travel; relevant exposures during travel (such as food and water sources, animal contact, insect bites, healthcare exposure, and sexual contacts); the interval between return and symptom onset; the duration of symptoms at presentation; and the total time elapsed since return (Figure 1A) (Thwaites & Day, 2017; Boggild et al., 2011). The relationship between return date and symptom onset is particularly informative: whether symptoms began before, during, or after travel constrains the range of plausible diagnoses given the incubation periods of relevant pathogens (Thwaites & Day, 2017). This temporal information, combined with destination-specific disease epidemiology, allows clinicians to stratify risk and prioritise investigations (Wilson et al., 2007). Systematically capturing these variables (tabulated in Section 4.2) could allow real-time identification of high risk patients and enable decision support around selection of appropriate panels of diagnostic tests. Applied across whole patient populations, it would enable surveillance systems to quantify the contribution of international travel to the burden of specific infections, identify outbreaks, and to track the importation of resistant organisms (Sridhar et al., 2021; Wilson et al., 2007), including those associated with travel to regions affected by natural disasters or conflict.

**Figure 1:**
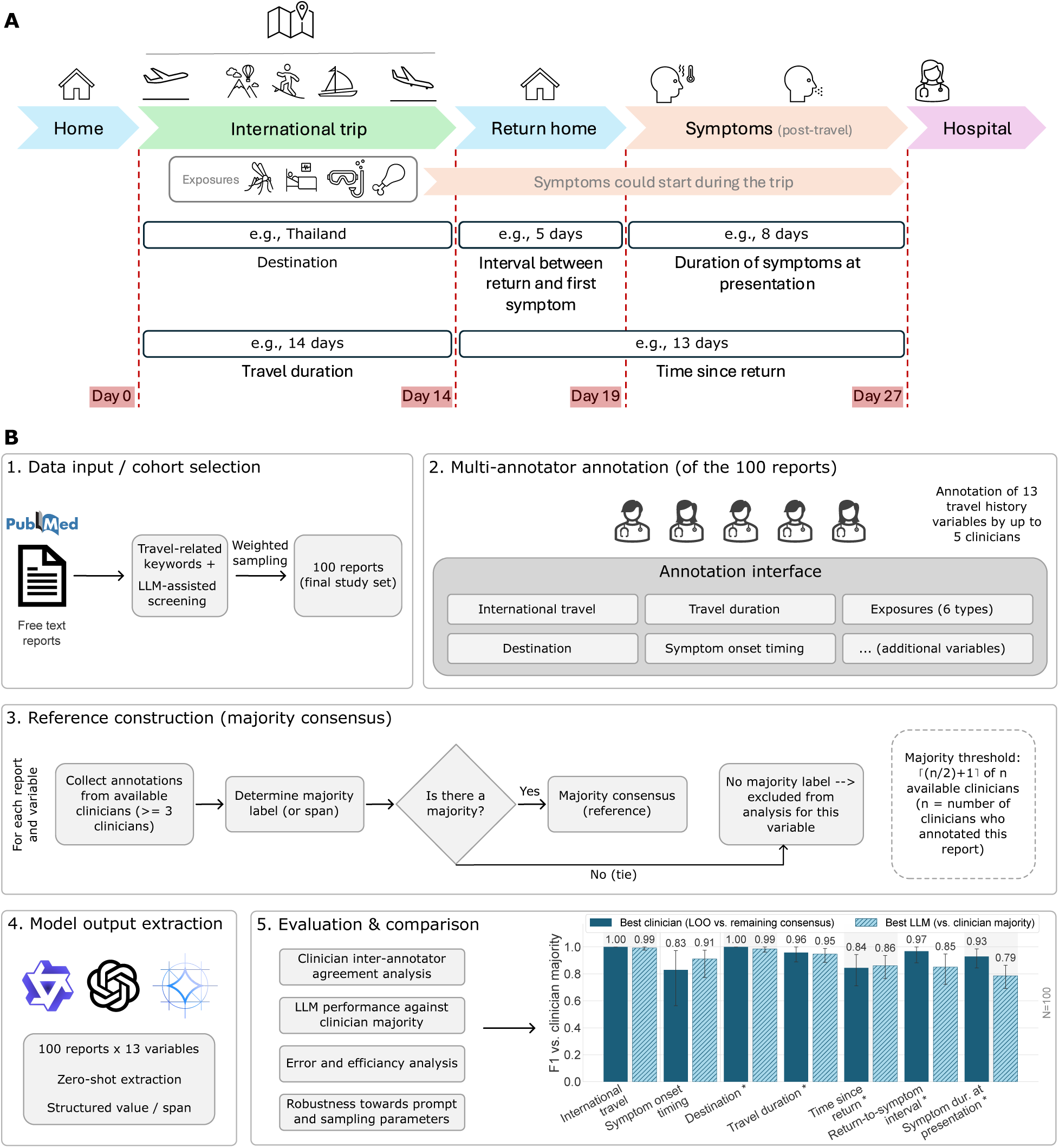
Study overview: travel history variables and annotation pipeline. **(A)** Illustrative timeline of a patient’s journey through international travel, showing key events, exposures that may occur during the trip, and the time-related milestones relevant to a travel-associated presentation. Day labels and durations shown are illustrative. **(B)** Pipeline: case reports were sampled via keyword filtering and LLM-assisted screening (1), and independently annotated by up to five clinicians across 13 travel history variables (2), aggregated into a per-note majority consensus with ties excluded (3). LLM outputs were extracted zero-shot in the same structured format (4) and compared against clinician annotations and consensus for agreement, accuracy, error patterns, and robustness (5). Bar chart shows F1 for the best-performing LLM against clinician consensus, alongside the best clinician’s leave-one-out F1; span variables (asterisked) reflect presence only.

Large language models (LLMs) have demonstrated considerable capability in extracting structured information from clinical text across a range of domains, including medication histories, diagnoses, and patient demographics (Lopez et al., 2025; Agrawal et al., 2022; Arsenyan et al., 2024). Their potential to extract travel-related clinical variables from free-text reports has not been systematically evaluated. Such evaluation is a necessary precondition for any deployment of LLM-based extraction in clinical settings, infectious disease surveillance, or AMR research pipelines, and requires a reliable reference standard against which model outputs can be assessed.

Unlike many established clinical information extraction tasks, travel history combines explicit entity extraction with temporal reasoning across multiple events. Relevant information is often distributed across the clinical narrative rather than expressed in a single sentence, requiring the relationship between travel, return, and symptom onset to be reconstructed from context. These characteristics make travel history a qualitatively different extraction problem from medications, diagnoses, or demographic attributes, for which benchmark datasets and evaluation protocols are already well established (Mahajan et al., 2023; Hu et al., 2026; Yang et al., 2019).

Here we describe a benchmark study in which we assess the accuracy of LLMs in extracting structured travel history variables from free-text clinical case reports. We developed an annotation framework covering 13 variables, including any travel mention (binary), destination, exposures, travel duration, symptom timing, and the temporal relationship between return and symptom onset. Five clinicians annotated a corpus of 100 case reports, yielding over 5,000 variable-level clinician annotations and providing a human reference standard against which LLM performance was measured. We report inter-annotator agreement, model accuracy relative to the clinician consensus, and the sources of systematic error. Our findings have implications for the practical feasibility of using LLMs to augment clinical practice, infectious disease surveillance and AMR monitoring through retrospective and prospective extraction of travel history from routine clinical documentation.

## 2 Results

To evaluate the feasibility of automated travel history extraction from clinical free text, we constructed an annotation dataset of 100 case reports from PMC-Patients-v2 (Zhao et al., 2023), annotated across 13 variables by five clinicians, yielding 5,174 variable-level clinician annotations (dataset construction: Section 4.1; annotation protocol: Section 4.3), and compared clinician and LLM extraction using a common pipeline (Figure 1B).

Published case reports differ from routine EHR documentation in that they are structured, editorially reviewed, and written retrospectively once the diagnosis is known (Zhang et al., 2025); we return to the implications of this for interpreting absolute performance in the Discussion. This setting nonetheless provides a controlled basis for comparing clinician and LLM extraction behaviour on the same material.

### 2.1 Clinician agreement is high for travel presence and destination but remains poor for temporal variables

We first characterised annotation consistency among clinicians before introducing any model outputs, across the 13 variables described in Section 4.2 (Table 1). Inter-annotator agreement varied substantially across variables (Figure 2A). The presence/absence of any sections of text (spans) identified as describing international travel, destination, and travel duration all reached reliable agreement (Krippendorff’s *α* = 0.95, 0.92, and 0.87 respectively). However, temporal variables in contrast showed relatively poor agreement: symptom onset timing and presence/absence of spans describing the remaining three temporal variables ranged from *α* = 0.53 to 0.59. As Krippendorff’s *α* is sensitive to category prevalence (Section 4.5), label distributions must be taken into account while interpreting the coefficients mentioned above (Appendix A.5, Figure 9).

**Figure 2:**
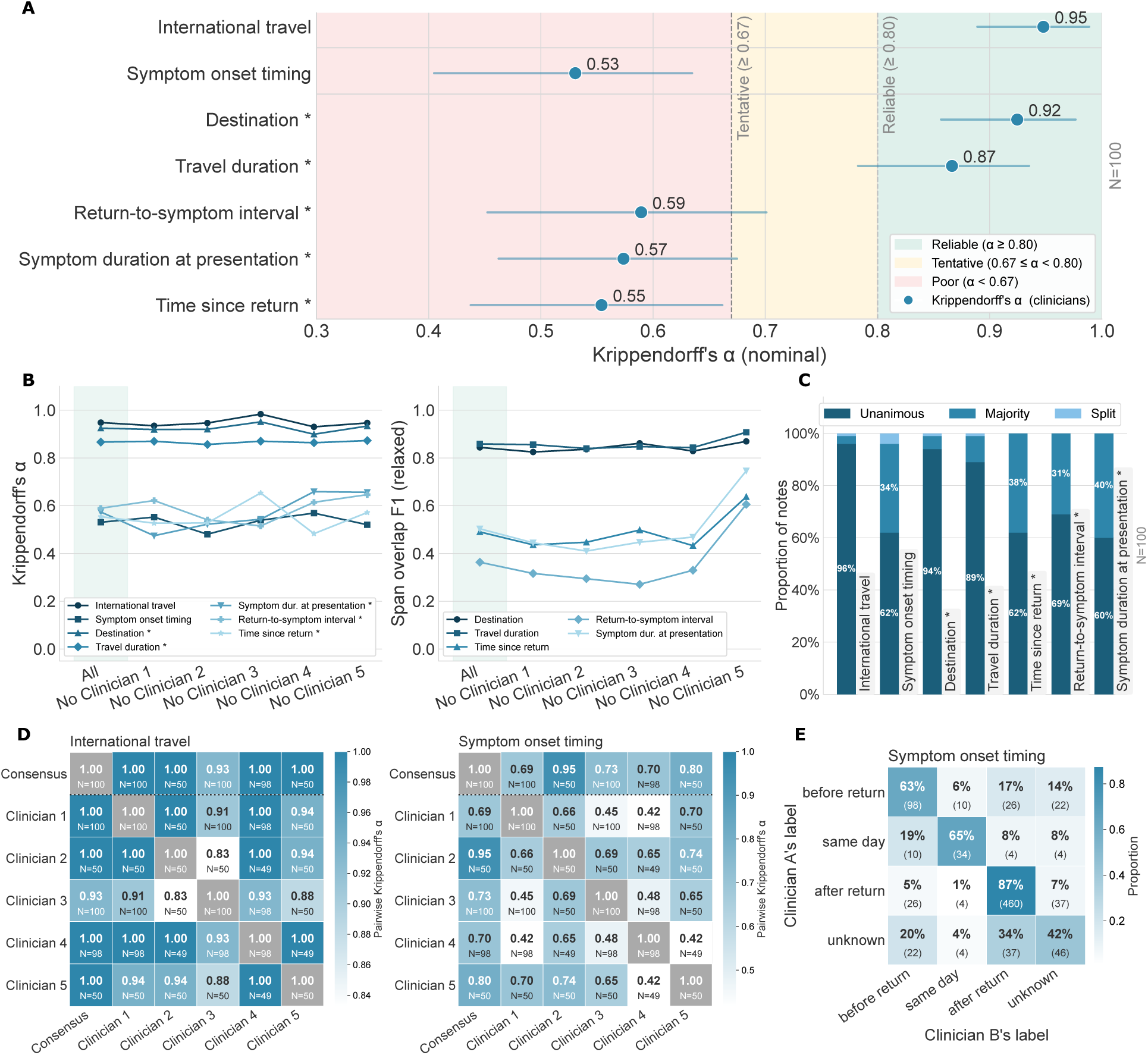
Inter-annotator agreement among clinicians across travel history variables. **(A)** Krip-pendorff’s *α* (nominal) across available clinician annotations for 100 notes per variable. For span variables (marked *), *α* reflects span presence only; span boundary agreement is reported separately. Whiskers: 95% bootstrap CI (N=2000 resamples). **(B)** Leave-one-out Krippendorff’s *α* for categorical variables (left) and relaxed span overlap F1 (Jaccard > 0.5) for span variables (right), recomputed after successively removing each clinician. Leftmost point: agreement across all available clinicians. **(C)** Proportion of notes with unanimous, majority, or split annotation outcomes per variable. Voting categories were determined among the clinicians who annotated each note; the number of contributing clinicians therefore varied by note. **(D)** Pairwise Krippendorff’s *α* heatmaps for international travel and symptom onset timing. The consensus row and column reflect agreement between the majority-vote label and each individual clinician. Cell counts may vary depending on overlapping notes. **(E)** Bidirectional label co-occurrence matrix for symptom on-set timing, pooled across all annotator pairs, showing the proportion and count of label assignments. All symptom-anchored variables (symptom onset timing, return-to-symptom interval, symptom duration at presentation) are defined with respect to the first reported symptom.

**Table 1:** Travel history variables annotated in this study.

| Variable | Definition | Label type |
| --- | --- | --- |
| International travel | Whether the clinical note states that the patient travelled internationally prior to presentation. | Binary |
| Destination | Country or geographic location visited during the reported international travel. | Span |
| Travel duration | Duration of the international trip. | Span |
| Time since return | Elapsed time between returning from travel and clinical presentation. | Span |
| Return-to-symptom interval | Time between travel return and onset of the first reported symptom. Symptoms could start before the return as well. | Span |
| Symptom duration at presentation | Duration of the first reported symptom at the time of clinical presentation. | Span |
| Symptom onset timing | Categorical variable indicating whether the first reported symptom began before travel, during travel, after return, or when timing could not be determined. | Categorical |
| Environmental exposure | Exposure to environmental sources relevant to infectious disease risk (e.g. freshwater or soil). | Categorical |
| Bites / animal exposure | Reported insect bites or contact with animals during travel. | Categorical |
| Food / water exposure | Exposure through food or drinking water during travel. | Categorical |
| Sick contacts | Reported contact with an ill individual during travel. | Categorical |
| Sexual contact | Reported sexual contact during travel. | Categorical |
| Healthcare exposure | Contact with healthcare facilities or medical procedures during travel. | Categorical |

This pattern held under leave-one-out analysis (Section 4.5): the agreement profile for categorical variables was largely stable regardless of which clinician was removed (Figure 2B). For span variables, removing one specific clinician improved overlap F1 for temporal spans, identifying that annotator as a partial outlier on those fields. The distribution of voting outcomes showed similar results (Figure 2C) with temporal variables produced split or majority outcomes in nearly 40% of cases, compared to near-unanimity for travel presence, destination, and duration. A qualitative comparison of disagreement sources for travel duration and time since return is given in Appendix A.7.

Pairwise agreement heatmaps for two representative variables illustrated the same result at the annotator level (Figure 2D; full set in Appendix A.6). For international travel, all annotator pairs showed uniformly high agreement; for symptom onset timing, pairwise scores were more variable, with label co-occurrence showing where disagreements were concentrated (Figure 2E).

### 2.2 Larger LLMs approach clinician-level performance on well-defined variables but struggle where clinicians do

We evaluated several LLMs (Appendix A.2) against the majority clinician consensus, using F1 for binary, categorical, and span presence variables, Matthews Correlation Coefficient (MCC) for exposure categories given label imbalance (Chicco & Jurman, 2020), and relaxed span overlap F1 for boundary assessment.

Performance tracked the difficulty pattern observed among clinicians (Figure 3A). International travel, destination, and travel duration were reliably extracted by most models, with scores clustering within or just below the clinician leave-one-out range. Temporal span variables and symptom onset timing showed wider dispersion in both clinician and model scores, consistent with these variables reflecting a possible ambiguity in the source text rather than a model-specific weakness. Smaller models, particularly qwen3.5-2b, clearly performed worse across most variables.

**Figure 3:**
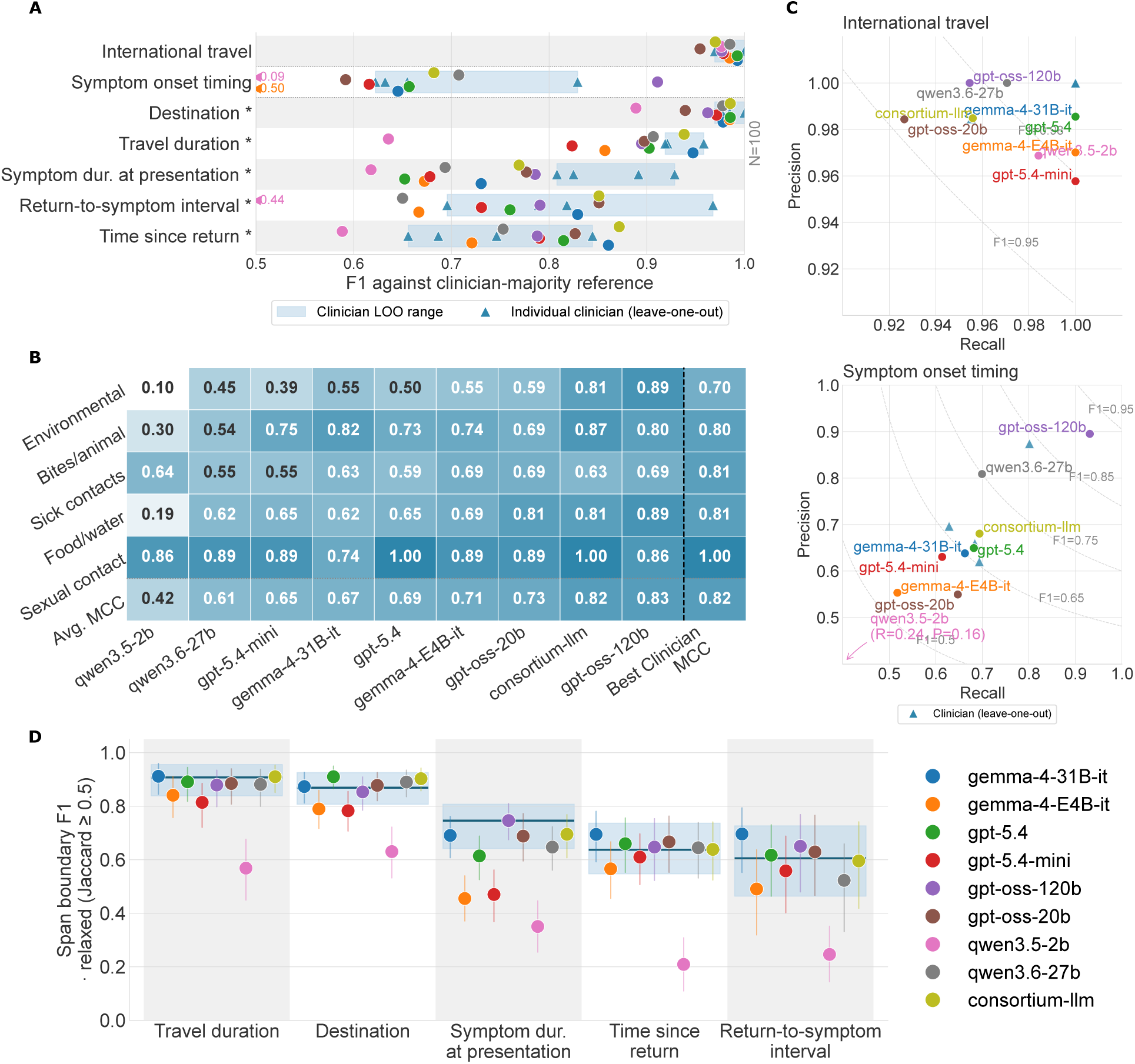
LLM performance against clinician majority consensus across travel history variables. **(A)** F1 score for each LLM (coloured circles) against the clinician majority consensus across binary, categorical, and span presence variables (N=100 per variable). Span variables (marked *) reflect span presence only; span boundary agreement is shown in D. The shaded band shows the clinician leave-one-out (LOO) F1 range; triangles show individual clinician LOO values. Clinician LOO estimates use the available overlapping annotations and therefore have clinician-specific sample sizes. **(B)** Matthews Correlation Coefficient (MCC) for each LLM against majority clinician labels for various exposure categories, used in preference to F1 given severe label imbalance. The rightmost column shows the best single-clinician MCC against majority consensus. **(C)** Precision-recall plots for international travel (top) and symptom onset timing (bottom), with clinician LOO values shown as triangles and iso-F1 contours overlaid. **(D)** Relaxed span boundary F1 (Jaccard *≥* 0.5) per LLM, computed by matching model spans against each clinician’s annotations individually and averaging across clinicians and cases. Solid line and shaded band show mean and 95% bootstrap CI of pairwise clinician span overlap, computed by treating each clinician in a pair alternately as prediction and reference. All symptom-anchored variables are defined with respect to the first reported symptom.

Exposure categories showed comparable overall performance between the strongest models and the best clinician (consortium-llm and gpt-oss-120b (Agarwal et al., 2025): average MCC 0.82 and 0.83; best clinician: 0.82), with substantial variation by exposure type, which was stronger for sexual contact, and weaker for environmental and food/water exposure (Figure 3B).

Precision-recall profiles (Figure 3C; full set in Appendix A.8, Figure 11) showed tight clustering near perfect precision and recall for international travel, but a more dispersed, model-specific distribution for symptom onset timing, without a consistent bias toward over- or under-prediction. For time since return and symptom duration at presentation specifically, smaller models tended to be recall-favoured relative to clinicians.

Span boundary agreement (relaxed Jaccard *≥* 0.5; Figure 3D), which reflects how consistently clinicians and models labelled specific sections of text followed the same ordering: larger models matched or approached the clinician pairwise mean for travel duration and destination, with performance declining for the three temporal spans. Under strict exact-offset matching (Appendix A.9, Figure 12), absolute F1 dropped across all models and variables while relative ordering was preserved. Decomposing destination error into boundary and label components under three progressively relaxed matching criteria (Figure 4; Appendix A.4) showed that F1 progressively increased when span boundaries were ignored and partial lexical matches were accepted (e.g., Kuwait City versus Kuwait), indicating that a substantial proportion of errors reflected boundary or lexical mismatches rather than wholly incorrect destination identification. Because many destination errors reflect boundary or granularity differences, destination extraction may be more usable for surveillance than span-level F1 alone suggests.

**Figure 4:**
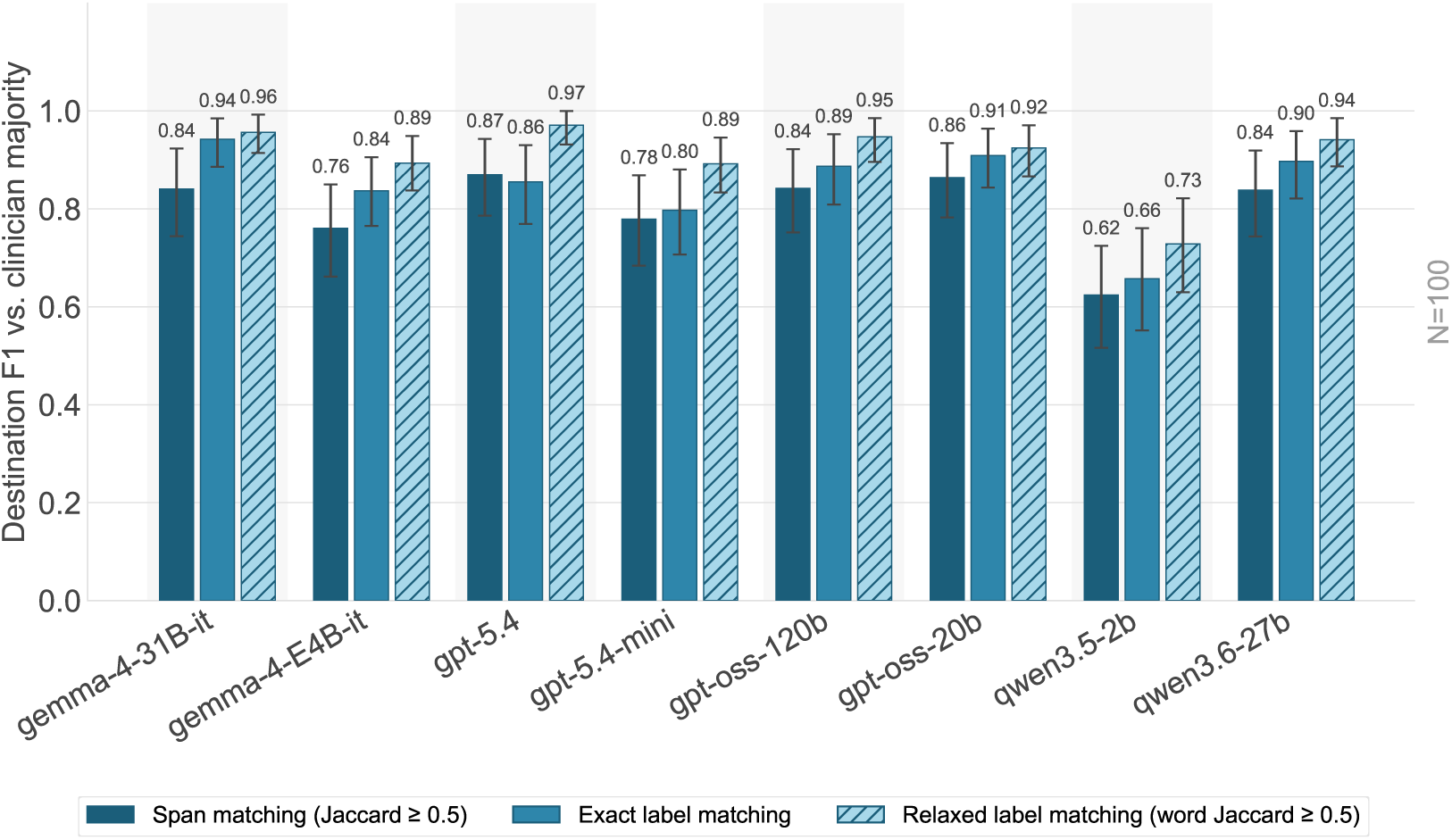
Destination extraction performance under progressively relaxed matching criteria. Performance is reported as F1 against the clinician-majority reference for several LLMs (N = 100 notes). Span matching requires character-offset Jaccard similarity *≥* 0.5 under greedy one-to-one assignment. Exact label matching ignores character positions and compares normalized destination text values by exact string equality. Relaxed label matching further accepts matches with word-level Jaccard similarity *≥* 0.5, accommodating differences in geographic granularity (e.g., Kuwait City versus Kuwait). Error bars denote 95% percentile bootstrap confidence intervals computed from 2,000 note-level resamples.

### 2.3 LLMs tend to make errors on cases where clinicians themselves disagree

To characterise where model errors occur, we ranked notes for each variable by clinician agreement, using the entropy of the available clinician labels for each note as a measure of case difficulty: low entropy indicating unanimous or near-unanimous agreement (easy), high entropy indicating substantial disagreement (hard). For each language model, we recorded whether its prediction matched the clinician majority label and computed a rolling error rate across notes ordered from easy to hard (Figure 5).

**Figure 5:**
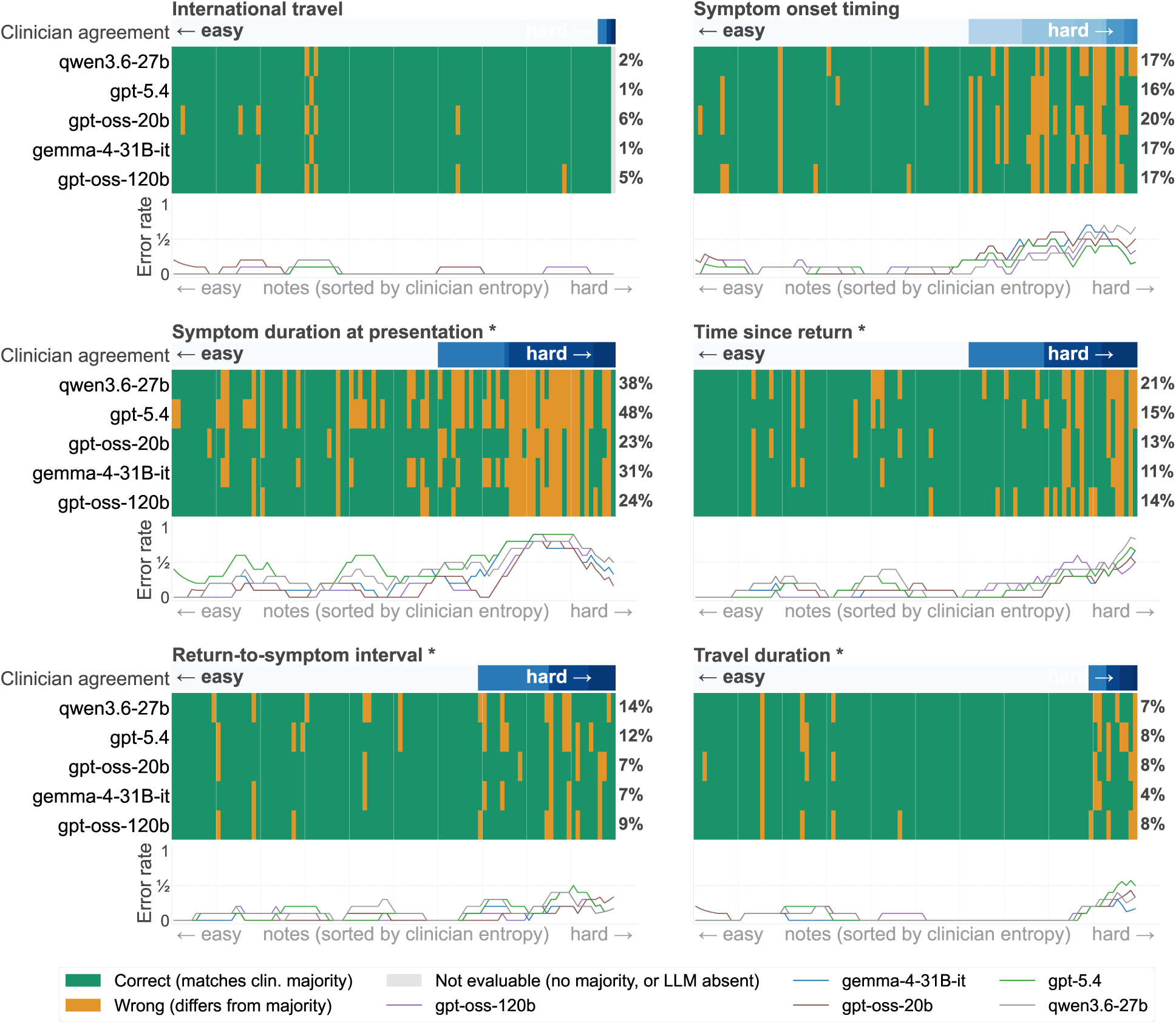
Distribution of LLM errors relative to clinician agreement. Each panel shows model performance for one variable, with notes (N=100) ordered left to right by clinician agreement, from unanimous (easy) to highly discordant (hard), measured as the entropy of the available clinician labels for each note. Top strip in each panel indicates clinician agreement level per note (darker shading: lower agreement). Heatmap rows show, for each model, whether its prediction matched the clinician majority label (green: correct; yellow: incorrect; grey: not evaluable, due to no clinician majority or model non-response). Right-hand percentages show each model’s overall error rate for that variable. Bottom line plots show rolling error rate (proportion incorrect within a 10-note window) across the same easy-to-hard ordering, by model. All symptom-anchored variables are defined with respect to the first reported symptom.

For most variables, model errors were concentrated toward the hard end of this ordering. Time since return and travel duration showed the clearest pattern: error rates remained low and flat across the easy and midrange notes, rising only once clinician disagreement became substantial. Thus, for these variables, model failures largely coincided with cases that are also difficult for clinicians, rather than reflecting independent model weaknesses.

Two variables departed from this pattern. For international travel, an easy task with high clinician agreement throughout, model errors were sparse but distributed across the full range of notes rather than concentrated at the hard end, indicating that the small number of errors made here are not explained by case difficulty.

Symptom duration at presentation showed elevated error rates even among easy notes, suggesting an additional source of difficulty for this variable beyond clinician disagreement, potentially related to how symptom duration is expressed in text independent of any ambiguity in onset timing itself.

### 2.4 LLM extraction is broadly robust to prompt wording but sensitive to decoding temperature

We assessed sensitivity to prompt wording, prompt comprehensiveness, and sampling temperature using the setup described in Section 4.6.

Performance was largely stable across five paraphrased prompt variants with fixed output structure (Figure 6A), though symptom onset timing, already the lowest-agreement variable among clinicians showed the most fluctuation. Self-consistency across prompt variants, measured with Krippendorff’s *α* on a larger sample (Figure 6B), exceeded the reliable threshold (*α* > 0.80) for all three models and variables, indicating that model outputs remained internally consistent despite case-level fluctuation. Similarly, reducing prompt comprehensiveness (Figure 6C, Figure 6D) had little effect on international travel, with a small variations on time since return and symptom onset timing. Gemma-4-31b-it (Team, 2026) was consistently the most stable model across both prompt manipulations; gpt-oss-120b the least.

**Figure 6:**
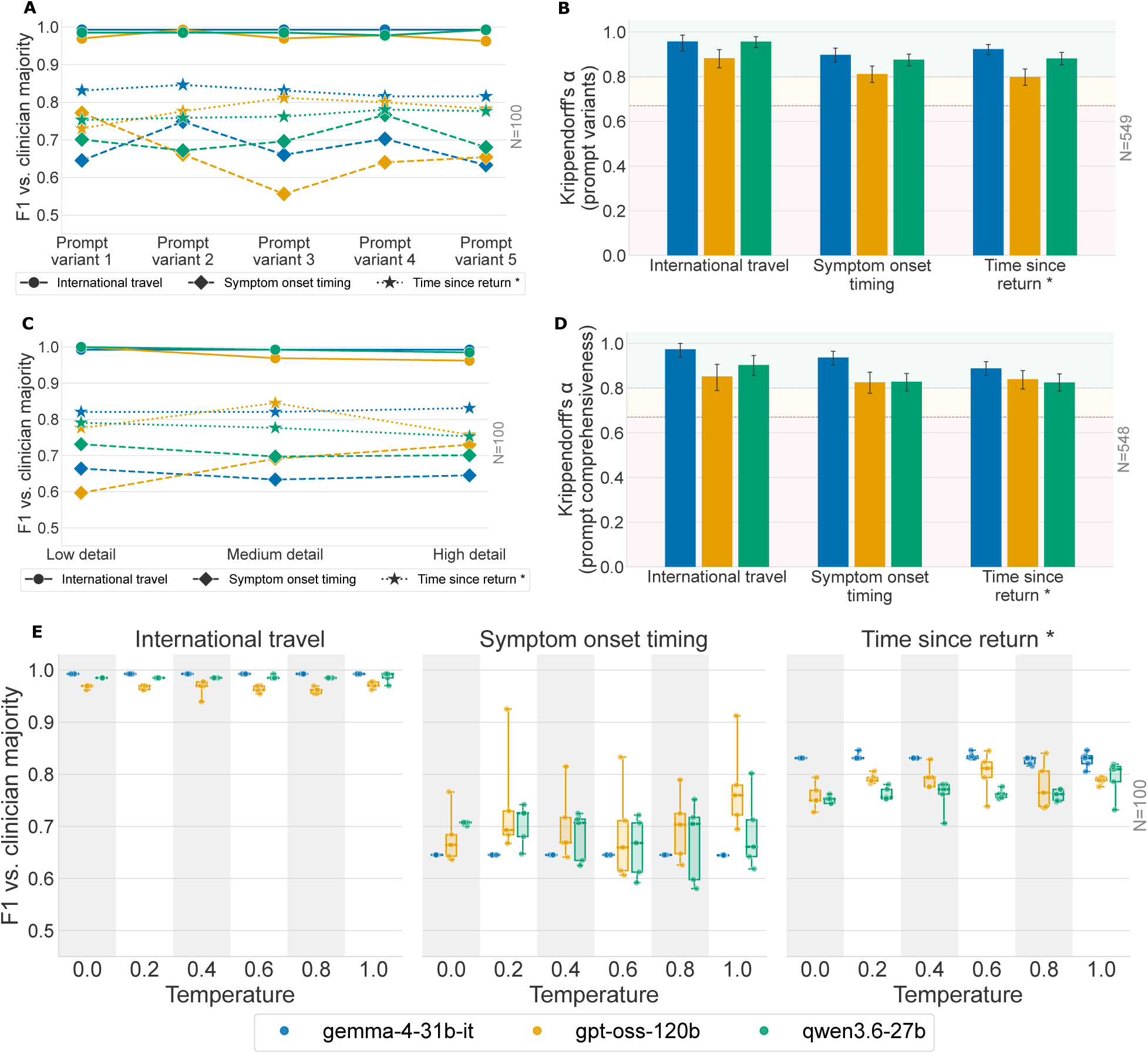
Sensitivity of LLM extraction to prompt and decoding variation. All results shown for three models (gemma-4-31b-it, gpt-oss-120b, qwen3.6-27b) and three representative variables (international travel, symptom onset timing, time since return), at temperature 0.0 unless otherwise noted. (A) F1 against clinician majority across five paraphrased prompt variants with fixed output structure (N=100). (B) Krippendorff’s alpha across the five prompt variants in A, computed on a larger random sample from the candidate pool (N=549). Error bars show 95% bootstrap CI. Dashed lines mark the tentative (*α* = 0.67) and reliable (*α* = 0.80) agreement thresholds. (C) F1 against clinician majority across three prompt comprehensiveness levels (low *≈* 45%, medium *≈* 70%, high = 100% of default token count), with output structure held constant (N=100). (D) Krippendorff’s alpha across the three comprehensiveness levels in C (N=548). (E) F1 against clinician majority across six sampling temperatures (0.0–1.0), five runs per temperature using the default prompt (N=100). Each point is one run; box plots show the median and interquartile range across the five runs per temperature.

Sampling temperature had a larger effect. International travel remained stable across the 0.0–1.0 range, but variance increased with temperature for symptom onset timing and time since return, across all three models (Figure 6E). Gpt-oss-120b showed the widest spread, including at temperature 0.0, which could be attributed to variability introduced during its reasoning/inference stack rather than sampling itself, a point we did not investigate further.

## 3 Discussion

### LLM performance approaches clinician agreement for explicit travel history variables but degrades for temporal inference

This study benchmarked LLM performance against a clinician-derived reference standard across binary, categorical, and span variables describing travel history in free-text clinical case reports. Three findings emerged. First, both clinicians and LLMs achieved high agreement for variables with explicit textual evidence, including international travel, destination, and travel duration, while performance declined for variables requiring temporal inference, particularly symptom onset timing. Second, LLM errors were concentrated on the same cases that produced clinician disagreement, suggesting that much of the remaining difficulty is intrinsic to the source text rather than specific to current models. Third, model outputs were considerably more sensitive to sampling temperature than to prompt wording or comprehensiveness, indicating that decoding configuration has a greater influence on extraction stability. Together, these findings indicate that LLM-based extraction is already feasible for a meaningful subset of travel history variables, including travel destination, which is particularly important for surveillance and epidemiological applications. However, the harder variables, focused on temporal features and most relevant to individual case management, require either improved modelling or a reconsideration of how they are defined and reported.

### Where LLMs and clinicians disagree

Variables that performed well were generally stated explicitly and required little inference. In contrast, temporal variables require reconstruction of a timeline from information that is often incomplete, distributed across the narrative, or only loosely expressed. This pattern is consistent with previous work showing that LLMs perform well on extraction tasks with clear lexical anchors but degrade on tasks requiring temporal or relational reasoning over clinical text (Kougia et al., 2024; Wang & Zhao, 2024; Zhang et al., 2024; Kruse et al., 2025). Importantly, clinicians showed the same pattern of disagreement (Figure 2A), and the structure of this disagreement closely mirrored LLM errors (Figure 5). Examination of individual cases suggests that some disagreement may stem from the annotation guidelines rather than pure textual ambiguity: annotators were instructed not to infer intervals not explicitly stated, so cases where the relevant duration was computable from other stated facts were scored as disagreement (Appendix A.7). Both sources may have contributed to the observed performance ceiling. Closing this gap will likely require both improved models and clearer reporting and annotation conventions.

### Toward deployment

Travel history is rarely captured as structured data within clinical systems (Chapman et al., 2010), limiting its use for surveillance and downstream analyses. Reliable extraction enables retrospective analyses of travel-associated infection and AMR acquisition at a scale that would be impractical through manual review (Peterson et al., 2021), while also supporting point-of-care tools that identify missing or incomplete travel histories during clinical documentation.

Successful deployment depends not only on extraction accuracy but also on practical constraints within healthcare systems. Many healthcare settings restrict transmission of clinical text to external services, motivating on-premises deployment of open-weight models (Jonnagaddala & Wong, 2025). Our results show that smaller open-weight models approach larger models on several explicit variables but remain weaker on temporal reasoning, suggesting that model selection should be guided by the variables of interest rather than overall benchmark performance. Temperature had a larger impact on extraction stability than prompt formulation, and reasoning-enabled models introduced substantial computational overhead, increasing inference latency and deployment cost (Al-Garadi et al., 2025). The throughput analysis in Appendix A.3 (Figure 8) illustrates this trade-off.

### Limitations

This study was conducted on published case reports rather than routine EHR notes. Case reports are more structured and consistently written than clinical documentation, and the reported performance should therefore be viewed as an upper bound for real-world deployment. This upper bound is likely a substantial one. Case reports are written retrospectively once the diagnosis is known, so the details included, or omitted, are shaped by that hindsight in a way prospective clinical notes are not, and they lack the noise typical of routine documentation. Performance here is therefore likely close to a ceiling: LLMs are trained more heavily on published biomedical text such as PubMed case reports than on real-world clinical notes, and the reports themselves are curated narratives rather than the messier text clinicians produce in practice. To our knowledge, this is the first benchmark comparing clinicians and LLMs on travel history extraction, but validation on genuine clinical notes remains essential. Clinical documentation also varies across institutions, which may limit generalisability.

The annotated dataset was intentionally enriched for travel-positive reports rather than sampled to reflect true prevalence. This increased the number of informative examples available for evaluating travel-related variables within limited clinician annotation time, but means prevalence-sensitive metrics should not be interpreted as representative of the source corpus (Section 4.1). Finally, several exposure categories contained relatively few positive examples, increasing uncertainty around the reported MCC estimates.

### Future directions

The most important next step is evaluation on real EHR data under appropriate governance, testing whether the same results of higher agreement on explicit variables and lower agreement on temporal variables generalises beyond published case reports. Public resources such as MIMIC (Johnson et al., 2023a) provide one possible starting point, although de-identification procedures often remove destinations, travel dates, and related attributes central to this task. Applying LLM extraction retrospectively to large clinical note collections would also provide a direct evaluation of the surveillance applications motivating this work.

Further progress will likely depend on improving extraction of temporal variables while also establishing clearer documentation and annotation conventions. More systematic evaluation of decoding strategies is similarly warranted, given their direct impact on the reliability and computational cost of large-scale clinical information extraction.

## Conclusion

Travel history remains an important but largely unstructured component of clinical documentation. Our results show that modern LLMs can reliably recover several clinically relevant travel history variables while also identifying the temporal relationships that continue to challenge both clinicians and models. These findings provide a practical foundation for integrating automated travel history extraction into infectious disease surveillance and future clinical decision support systems.

## 4 Methods

### 4.1 Dataset construction

The clinical case reports used in this study were drawn from PMC-Patients-v2 (Zhao et al., 2023), a collection of approximately 250,000 published case reports extracted from PubMed Central. To identify reports likely to contain travel history, we first applied a keyword-based filter using travel-related terms, yielding approximately 9,000 candidate reports. The keyword list is provided in the accompanying code repository.

Because travel history remains relatively uncommon even within this filtered subset, we further applied gpt-oss-120b (Agarwal et al., 2025) as a candidate selection model to identify reports describing international travel involving a single destination, producing approximately 1,300 travel-positive case reports suitable for downstream annotation. The final annotation set of 100 reports was constructed by randomly sampling from both the travel-positive and travel-negative pools, with approximately 70% drawn from the travel-positive pool and the remaining 30% from travel-negative reports. This enrichment strategy concentrated limited clinician annotation effort on informative cases; as a result, label prevalence in the annotated dataset does not reflect true prevalence in the source corpus, and prevalence-sensitive metrics should be interpreted accordingly (see Limitations).

As gpt-oss-120b was also included among the models evaluated in this study, its use for candidate selection raises a question of unfair advantage. However, its role at this stage was restricted to a coarse binary judgement of whether international travel involving a single destination was described, distinct from the full set of variables assessed against clinician consensus in the main evaluation. We do not consider this restricted, upstream use likely to advantage gpt-oss-120b on the downstream extraction task.

### 4.2 Annotation framework

We developed an annotation framework covering 13 travel history variables, covering binary (international travel), span (destination, travel duration, time since return, return-to-symptom interval, symptom duration at presentation), and categorical (symptom onset timing, six exposure categories) types, summarised in Table 1. All symptom-anchored variables were defined with respect to the first reported symptom where multiple symptoms were mentioned.

Both the annotation framework and the annotation application used to collect clinician labels were developed in-house. Prior to the main annotation round, the framework and variable definitions were presented to the clinician panel for review; queries and feedback from this session were incorporated into the final version of the guidelines used for annotation. Full annotation guidelines and a screenshot of the annotation interface are provided in Appendix A.1.

### 4.3 Clinician annotation protocol

Five clinicians independently annotated the case report corpus using an overlapping annotation design. Two clinicians annotated all 100 reports, one annotated 98, and two annotated 50 reports each, with assignments overlapping such that almost every report received annotations from at least three clinicians. The panel included an emergency medicine specialist trainee with five years of UK clinical practice, a senior infectious diseases registrar with over ten years of experience including work in high infectious disease burden settings and a Diploma in Tropical Medicine and Hygiene, and an infectious diseases trainee with eight years of clinical practice and postgraduate training in biomedical informatics and tropical medicine, and a licensed physician in China specialising in gastroenterology, whose training has been primarily research-based, and a UK-qualified doctor in Internal Medicine Training with three years of post-graduate experience and an NIHR Academic Clinical Fellowship. All annotators were DPhil students at the University of Oxford at the time of annotation, at various stages of doctoral research on applications of machine learning in infectious diseases and AMR.

Clinicians had access to the annotation guidelines (Appendix A.1) throughout annotation. Annotation was blinded: clinicians could not see other annotators’ labels at any point during the process. Consensus labels for each report were derived from the clinician annotations available for that report. For binary and categorical variables, the majority label among contributing clinicians was taken as consensus. This applied uniformly regardless of the number of possible labels: for symptom onset timing (four categories) and each of the six exposure categories (four response options: Yes, Denied, Unclear, Not mentioned), consensus was the label held by more than half of contributing clinicians, with no majority resulting in the note being excluded from that variable’s analysis. For span variables, overlapping clinician spans were clustered left-to-right, and a cluster was retained as consensus if it contained annotations from more than half of the clinicians who annotated that note (see Appendix A.4 for the full span-clustering procedure, applied here to destination but used identically for all span variables). Split votes, where no majority existed among contributing clinicians, occurred in a relatively small proportion of notes per variable (Figure 2C); these notes were excluded from the corresponding variable-level analysis where no majority could be determined.

### 4.4 Evaluated language models

We evaluated nine LLMs spanning proprietary API-served and open-weight model families, summarised in Appendix A.2 (Table 2). Open-weight models were served using vLLM. consortium-llm denotes a majority-vote consensus across five of the individual models (gpt-5.4, gpt-oss-120b, gpt-oss-20b, gemma-4-31b-it, qwen3.6-27b). Unless otherwise stated, models were run with default inference settings. Prompt templates and output schema are provided in the accompanying code repository.

### 4.5 Evaluation metrics

#### Inter-annotator agreement

Agreement among clinicians was quantified using Krippendorff’s *α* (nominal) (Krippendorff, 2011), defined as *α* = 1 *− D_o_/D_e_*, where *D_o_* is the observed disagreement across all annotator pairs and *D_e_* is the disagreement expected under chance, estimated from the observed marginal label distribution. Because *D_e_* depends on category prevalence, *α* is not directly comparable across variables with differing label balance; label distributions for all variables are reported in Appendix A.5 (Figure 9). We adopted conventional thresholds of *α ≥* 0.80 (reliable) and *α <* 0.67 (poor) for interpretation (Krippendorff, 2011).

For span variables, *α* was computed on span presence/absence only, i.e. whether a clinician identified any text for that field in a given note, independent of the exact boundaries selected. Boundary-level agreement was assessed separately using a relaxed Jaccard overlap criterion (coefficient *>* 0.5) between annotators’ selected spans, reported as span overlap F1.

Leave-one-out (LOO) agreement was computed by excluding one clinician at a time and recomputing *α* (categorical variables) or overlap F1 (span variables) across the remaining annotators, repeated for each of the five clinicians in turn. This isolated whether agreement was driven by a subset of annotators rather than reflecting consistent difficulty across the full panel (Figure 2B). Individual clinician LOO performance, reported alongside LLM results in Figure 3A, was computed analogously: each clinician’s annotations were compared against the majority consensus of the remaining clinicians for that note. Excluding one clinician could result in tied votes for some notes; therefore, performance was calculated only for notes where a majority label could be determined among the remaining clinicians.

#### LLM performance

LLM outputs were compared against the clinician majority consensus using F1 for binary, categorical, and span presence variables; Matthews Correlation Coefficient (MCC) for the six exposure categories, given severe label imbalance (Chicco & Jurman, 2020); and relaxed span overlap F1 (Jaccard *≥* 0.5) for span boundary evaluation. As LLMs generate entity text rather than explicit character offsets, predicted text was post-processed into character spans prior to evaluation (Semin et al., 2026; Abdul et al., 2024). We additionally reported exact-offset (strict) span matching in Appendix A.9 as a lower bound alongside the relaxed criterion.

For destination specifically, we further decomposed span-level error into boundary and label components using three progressively relaxed matching criteria: span matching (character-offset Jaccard *≥* 0.5, greedy one-to-one assignment), exact label matching (normalised destination text, exact string equality), and relaxed label matching (normalised destination text, word-level Jaccard *≥* 0.5). Full details of the clinician-majority reference construction under each criterion are given in Appendix A.4.

#### Case difficulty and error analysis

To characterise where model errors occurred, we ranked notes per variable by the entropy of available clinician labels, using this as a proxy for case-level annotation difficulty, and computed rolling model error rate across this easy-to-hard ordering (Section 2.3).

All confidence intervals reported for agreement, F1, and MCC estimates were computed via percentile bootstrap (2,000 resamples), with resampling performed at the note level unless otherwise specified.

### 4.6 Prompt and decoding sensitivity analysis

We assessed sensitivity to prompt wording, prompt comprehensiveness, and sampling temperature using three models drawn from different model families (gemma-4-31b-it, gpt-oss-120b, qwen3.6-27b), to isolate effects specific to any one training stack. Analyses were restricted to three representative variables spanning the difficulty range established in Section 2.2: international travel, symptom onset timing, and time since return.

Reasoning was disabled for gemma-4-31b-it and qwen3.6-27b due to generation time; gpt-oss-120b was run at its default (medium) reasoning effort throughout, as effort cannot be disabled for this model. Unless otherwise stated, all sensitivity analyses were run at sampling temperature 0.0.

Prompt wording robustness was assessed using five paraphrased prompt variants with fixed output structure. Prompt comprehensiveness was varied across three levels (low *≈* 45%, medium *≈* 70%, high = 100% of default token count) with output structure held constant. Temperature robustness was assessed by running each model five times at each of six temperature settings (0.0–1.0) using the default prompt. Prompt variants and comprehensiveness levels are provided in the accompanying code repository.

### 4.7 Inference infrastructure

Open-weight models were served on two NVIDIA RTX PRO 6000 Blackwell GPUs (96 GB VRAM each) using tensor parallelism via vLLM. Throughput and latency profiling for four representative open-weight models is reported in Appendix A.3 (Figure 8), using reasoning-enabled inference to characterise the computational overhead of chain-of-thought generation; this differs from the main experiments, which disable reasoning for gemma and qwen.

## Code and data availability

- Clinician annotated data along with LLM responses: https://hf.co/datasets/pkanithi/ travel-history-iaa-benchmark
- Prompts, IAA analysis and benchmark against LLMs: https://github.com/praveenkanithi/ travel-history-iaa-analysis
- PMC-Patients-v2: https://hf.co/datasets/zhengyun21/PMC-Patients

## Acknowledgment

This study is supported by the National Institute for Health and Care Research (NIHR) Health Protection Research Unit: Healthcare Associated Infections and Antimicrobial Resistance (NIHR207397), a partnership between the UK Health Security Agency (UKHSA) and the University of Oxford, and by the Oxford NIHR Biomedical Research Centre. The views expressed are those of the authors and not necessarily those of the NIHR, UKHSA or the Department of Health and Social Care. ASW is an NIHR Senior Investigator.

### A Appendix

#### A.1 Annotation guidelines and interface

Full annotation guidelines provided to clinicians, reproduced verbatim below. Note that two exposure categories described in the guidelines, *Percutaneous* and *Healthcare Exposure Abroad*, are not reported in the main text or figures, as no cases of either were observed in the annotated dataset.

**Figure 7:**
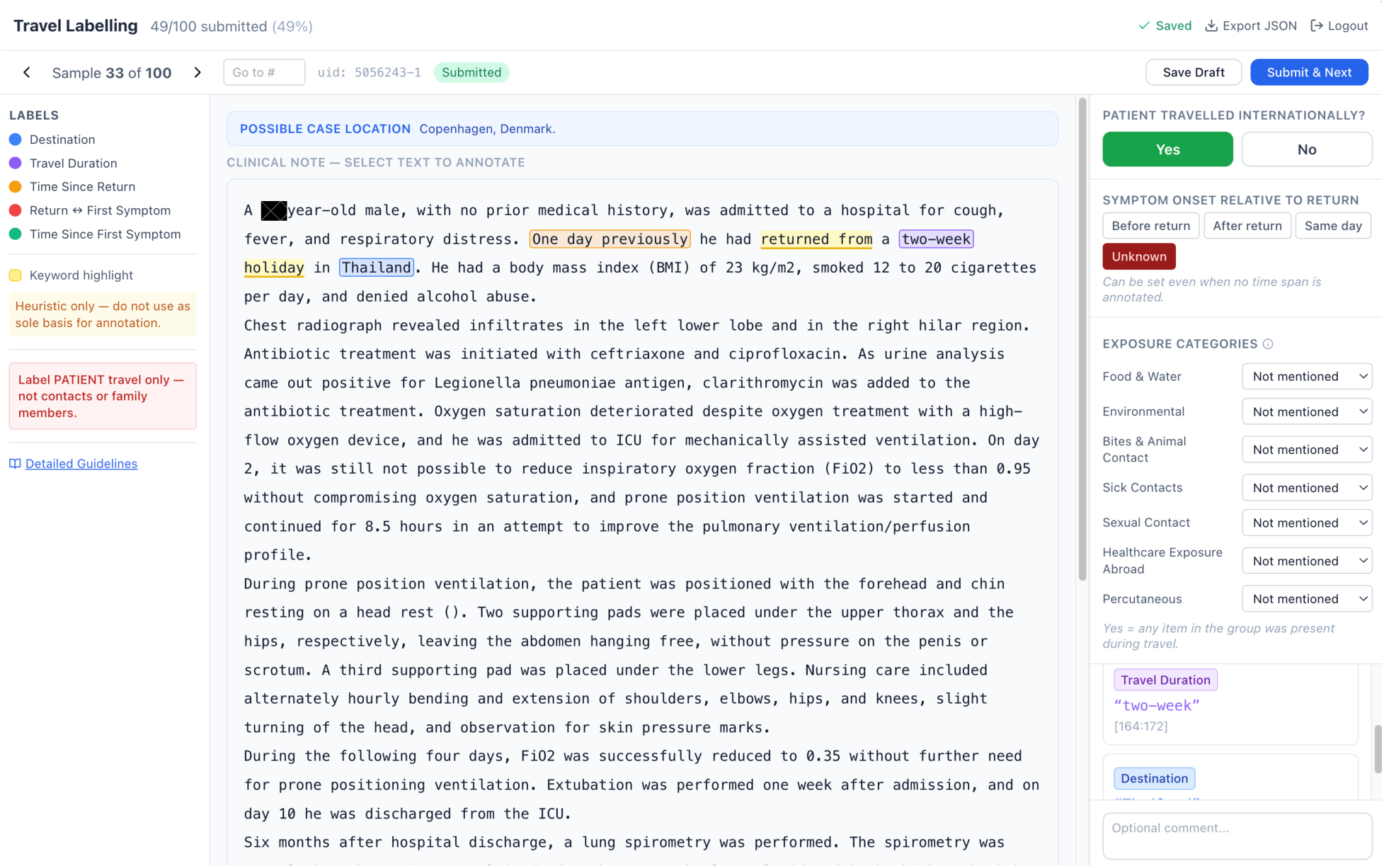
Annotation application interface. Screenshot of the in-house annotation tool used by clinicians to label travel history variables.

##### Guidelines

**Important:** Label only information about the patient’s travel — not contacts, relatives, household members, or other people.

###### Destination

Country or specific location of travel. Label country if mentioned; otherwise label city/region. Skip if only ‘overseas’/‘abroad’.

###### Travel Duration

Duration of the trip. Select only the time expression itself.

###### Time Since Return

Time from return from travel to hospital presentation. Keep span concise.

###### Return *↔* First Symptom

The explicit time gap between return and first symptom onset. Refers to the first reported symptom, regardless of whether it was the presenting complaint. Valid whether onset was before or after return. Leave blank if no explicit gap is stated.

###### Symptom Duration at Presentation

Time from first symptom onset to admission/presentation. Refers to the first reported symptom, regardless of whether it was the presenting complaint. May be available regardless of travel.

###### Return *↔* First Symptom Timing

Two independent fields capture the relationship between return and first symptom onset:

- Use Return *↔* First Symptom (span) when the note explicitly states a time gap — e.g. “8 days”, “two days”. Valid whether onset was before or after return.
- Use Symptom onset relative to return (categorical) to indicate direction: Before return, After return, Same day, or Unknown. This field may be inferred from context even when no explicit time gap is stated — e.g. “the fever started before returning home”.
- Both fields refer to the first reported symptom, regardless of whether it was the presenting complaint.
- Both fields are optional if the note does not make this clear.

###### Time-Span Rules

- All time spans must be explicitly stated in the note — do not infer or calculate timing.
- Label the time expression itself, including quantitative modifiers that form part of it (“more than 2 weeks”, “approximately 3 months”, “at least 5 days”). Exclude framing phrases that are not part of the duration (“before admission”, “prior to hospitalization”, “ago”).
- Ranges and compound durations are a single span (“2 to 3 weeks”, “2 weeks and 3 days”).
- Vague expressions like “few days”, “few weeks”, or “several weeks” are valid spans.
- Absolute date intervals are valid if that’s how timing is expressed.

###### Destination Rules

- If a country is mentioned, label the country.
- If only a city, island, or region is mentioned, label that instead.
- If city and country are mentioned together, label only the country.
- If the destination is only “overseas” or “abroad”, do not label destination.

###### Exposure Categories (travel only)

Seven exposure groups are assessed for each sample where international travel is confirmed. Each group has four possible responses:

- Yes
- Denied
- Unclear
- Not mentioned

Scoring rule: Mark Yes for a group if any single item within that group was present or the patient was exposed to it during travel. Absence of all items = Denied. Use Unclear when the note is ambiguous. Use Not mentioned when the note gives no information on the group.

The items listed under each group are examples of common exposures within that broad category. Any clinically similar exposure should be considered under the appropriate group.

###### Food & Water

- Tap or untreated water
- Unpasteurised dairy
- Raw or undercooked meat or seafood
- Raw produce

###### Environmental

- Freshwater contact
- Soil contact

###### Bites & Animal Contact

- Mosquito bite
- Tick bite
- Other insect or arthropod bite
- Animal bite or scratch
- Mammal contact
- Bird contact

###### Sick Contacts

- Contact with anyone ill during or after travel (e.g. family, friends, fellow travellers)

###### Sexual Contact

- New partner, unprotected, commercial, or multiple partners

###### Healthcare Exposure Abroad

- Medical injections, transfusions, procedures, or hospitalisations abroad

###### Percutaneous

- Tattoos or piercings
- IV drug use

###### Keyword Highlighting

Caution: Yellow-highlighted keywords are heuristic aids only. They do not determine the annotation. Always base your annotation on the full meaning of the note.

#### A.2 Evaluated language models

Table 2 summarises the language models evaluated in this work together with their source repositories. Unless otherwise stated, models were used with their default inference settings described in the main text.

**Table 2:** Language models evaluated in this study.

| Model | Type | Source |
| --- | --- | --- |
| gpt-5.4 | API | OpenAI |
| gpt-5.4-mini | API | OpenAI |
| gpt-oss-120b | Open-weight | https://hf.co/openai/gpt-oss-120b |
| gpt-oss-20b | Open-weight | https://hf.co/openai/gpt-oss-20b |
| gemma-4-31b-it | Open-weight | https://hf.co/google/gemma-4-31B-it |
| gemma-4-E4B-it | Open-weight | https://hf.co/google/gemma-4-E4B-it |
| qwen3.6-27b | Open-weight | https://hf.co/Qwen/Qwen3.6-27B |
| qwen3.5-2b | Open-weight | https://hf.co/Qwen/Qwen3.5-2B |
| consortium-llm | Open-weight | Consensus of five LLMs using majority voting: gpt-5.4, gpt-oss-120b, gpt-oss-20b, gemma-4-31b-it, qwen3.6-27b |

### A.3 Inference cost and throughput of reasoning LLMs

**Figure 8:**
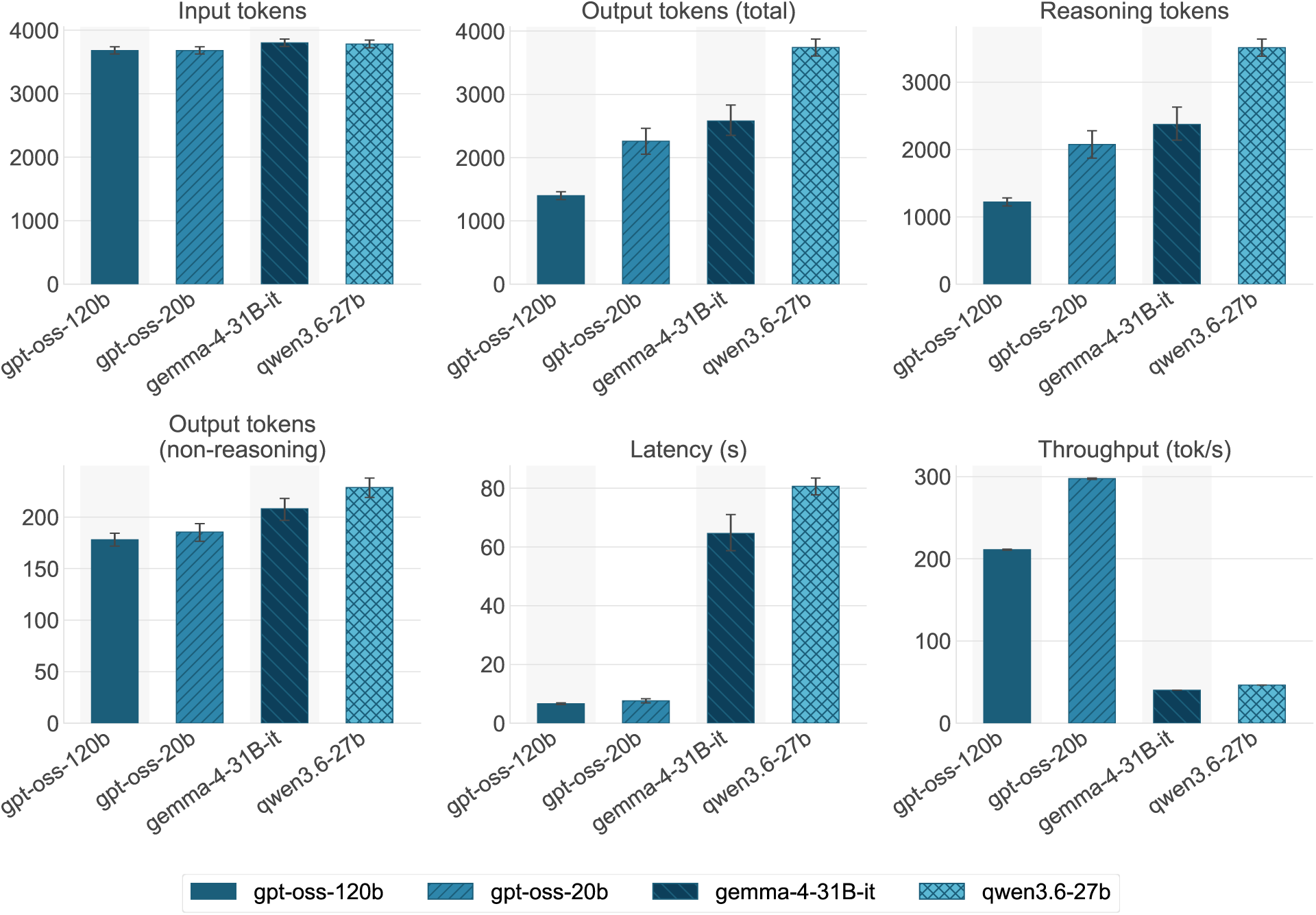
Inference cost and throughput of reasoning-enabled LLMs. Profiling results for four open-weight models evaluated on 150 randomly selected clinical notes using temperature = 0.0. All models were run sequentially on two NVIDIA RTX PRO 6000 Blackwell GPUs (96 GB VRAM each) with tensor parallelism. Panels report mean input tokens, total output tokens, reasoning tokens, non-reasoning output tokens, wall-clock latency, and throughput (total output tokens divided by latency). GPT-OSS models were evaluated using medium reasoning effort, while Gemma and Qwen were evaluated with reasoning enabled. Error bars denote 95% percentile bootstrap confidence intervals computed from 2,000 inference-level resamples.

Extraction accuracy is only one consideration when selecting an LLM for clinical information extraction. Practical deployment also depends on inference efficiency, including latency, throughput, and token usage. To characterize these computational costs, we profiled a representative subset of four open-weight reasoning models: gpt-oss-120b, gpt-oss-20b, gemma-4-31B-it, and qwen3.6-27b.

Unlike the main experiments in this manuscript, which disabled reasoning for Gemma and Qwen, this analysis evaluated all models with reasoning enabled to quantify the computational overhead associated with chain-of-thought generation. Gpt-oss models expose configurable reasoning effort (low, medium, and high); throughout this manuscript the reasoning effort was fixed to medium, including this additional experiment. In contrast, Gemma and Qwen provide only a binary reasoning mode (enabled or disabled) without configurable reasoning effort. Reasoning was enabled in the additional experiments reported here solely for profiling purposes and disabled in all other experiments (including the main experiments) because of the increased inference cost demonstrated below.

#### Experimental setup

Profiling was performed on 150 randomly selected clinical notes, which did not necessarily overlap with the 100 reports annotated by clinicians. Each model processed the notes sequentially. All models were served on identical hardware consisting of two NVIDIA RTX PRO 6000 Blackwell GPUs (96 GB VRAM each) using tensor parallelism. Temperature was fixed at 0.0 for all inferences.

For each inference we recorded six metrics: input token count, total output token count, reasoning token count, non-reasoning output token count, wall-clock latency (seconds), and throughput (output tokens per second, computed as total output tokens divided by latency). Confidence intervals were estimated using percentile bootstrap with 2,000 inference-level resamples.

#### Results

Input token counts were comparable across models (approximately 3,700–3,800 tokens per note; Figure 8). Although every model received the same prompt and clinical note, the reported token counts differ slightly because each model family employs its own tokenizer and vocabulary.

Output token usage varied substantially. Gpt-oss models generated approximately 1,400–2,300 output tokens per inference, whereas Gemma and Qwen generated approximately 2,600 and 3,700 tokens, respectively. These differences were driven primarily by reasoning token generation: Gemma and Qwen produced more than 2,000 reasoning tokens per inference on average, while gpt-oss generated substantially fewer under the medium reasoning setting. In contrast, the non-reasoning output was similar across all models (approximately 180–230 tokens per inference), reflecting the common structured JSON annotation format produced by every model.

These differences in reasoning token generation translated directly into inference latency. Gpt-oss-120b and gpt-oss-20b completed inference in a mean of 6.6 s (95% CI 6.3–6.9) and 7.6 s (6.9–8.3), respectively, achieving mean throughputs of approximately 211 and 298 output tokens/s. Gemma-4-31B-it required 64.6 s (58.7–71.0) and qwen3.6-27b required 80.6 s (77.7–83.5) per inference, with throughputs of approximately 40 and 46 output tokens/s, respectively.

The computational differences are consistent with the underlying model architectures. Gpt-oss-120b and gpt-oss-20b are mixture-of-experts (MoE) models with approximately 5.13B and 3.61B activated parameters per token, respectively, whereas gemma-4-31B-it and qwen3.6-27B are dense transformer models. Under otherwise identical hardware and decoding settings, the MoE models therefore achieve substantially lower inference latency and higher throughput than the dense models. These profiling results characterize the computational requirements of the evaluated implementations and are independent of the extraction accuracy comparisons presented in the main text.

### A.4 Destination extraction: matching criteria detail

Figure 4 in the main text reports destination extraction performance under three progressively relaxed matching criteria. This section provides the full definitions and reference construction underlying that comparison.

LLMs do not directly predict character offsets; instead, they generate destination text verbatim from the clinical note. As in most information extraction benchmarks, these generated text spans were post-processed into character spans and evaluated using span-based matching (Semin et al., 2026). While this provides a standard evaluation protocol, span-based metrics can conflate two qualitatively different error types: failing to localize the exact entity boundaries and failing to identify the correct destination itself.

#### Matching criteria

All evaluations used the same clinician-majority reference constructed from the available clinician annotations for each note. For span-based evaluation, overlapping clinician spans were clustered left-to-right, and a cluster was retained if it contained annotations from more than half of the clinicians who annotated the note. For label-based evaluation, clinician destination text values were normalized (lowercased and stripped), and a text label was included in the majority reference if it was produced by more than half of the annotators. This label majority was computed independently of the span clusters to avoid propagating boundary decisions into the label-based analyses.

- Span F1 required character-offset overlap between predicted and reference spans, with matches determined using greedy maximum-overlap assignment and accepted when the character-level Jaccard similarity is at least 0.5.
- Exact label F1 ignored character positions and instead compared normalized destination text values using exact string equality with one-to-one set matching.
- Relaxed label F1 further allowed partial lexical agreement by matching normalized destination labels when their word-level Jaccard similarity was at least 0.5 under greedy one-to-one assignment. This accommodated common granularity differences such as Kuwait City versus Kuwait or Sub-Saharan Africa versus Africa.

For all three metrics, 95% confidence intervals were estimated using percentile bootstrap with 2,000 note-level resamples.

### A.5 Label distributions across annotated variables

Presents the underlying label distributions (Figure 9) behind the Krippendorff’s *α* values reported in subsection 2.1, to provide an assessment of how skew in the label distribution may inflate or deflate chance-corrected agreement. Variables such as international travel, destination and time interval between return and the first symptom were slightly imbalanced, and hence *α* should be interpreted alongside these base rates rather than in isolation.

**Figure 9:**
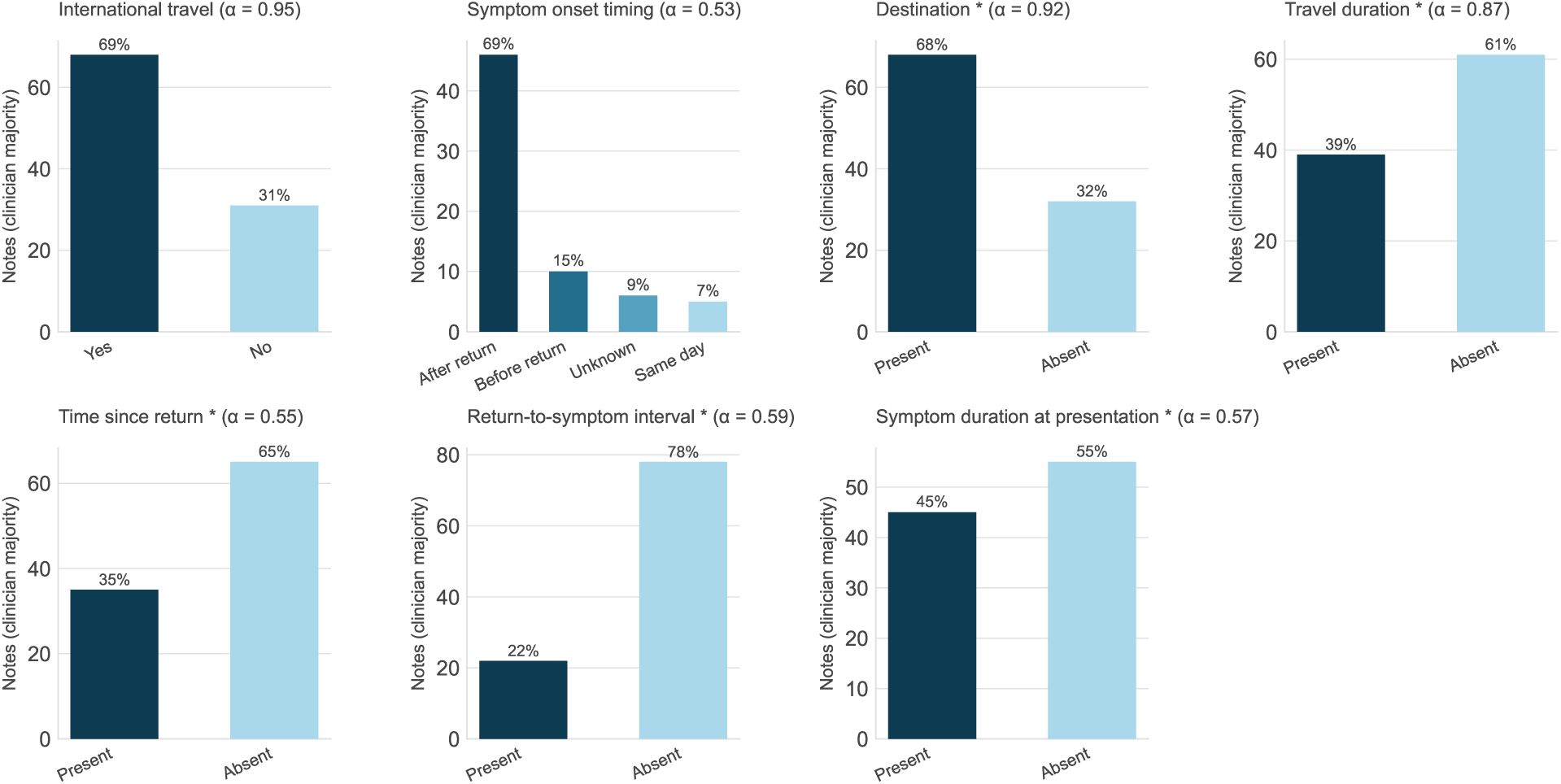
Data distribution of all variables. Bar charts showing the proportion of notes falling into each label category, based on the majority vote among available clinician annotations for each note (N=100 notes per variable). Binary and span-presence variables show the proportion of notes coded present/absent; symptom onset timing shows the proportion across its four categories. Krippendorff’s *α* (from Figure 2A) is reported in each panel title for reference against the underlying label distribution.

### A.6 Pairwise clinician agreement across all variables

Extends the two representative pairwise agreement heatmaps in Figure 2D (international travel, symptom onset timing) to all seven variables, giving the full clinician-by-clinician and consensus-by-clinician breakdown (Figure 10)

**Figure 10:**
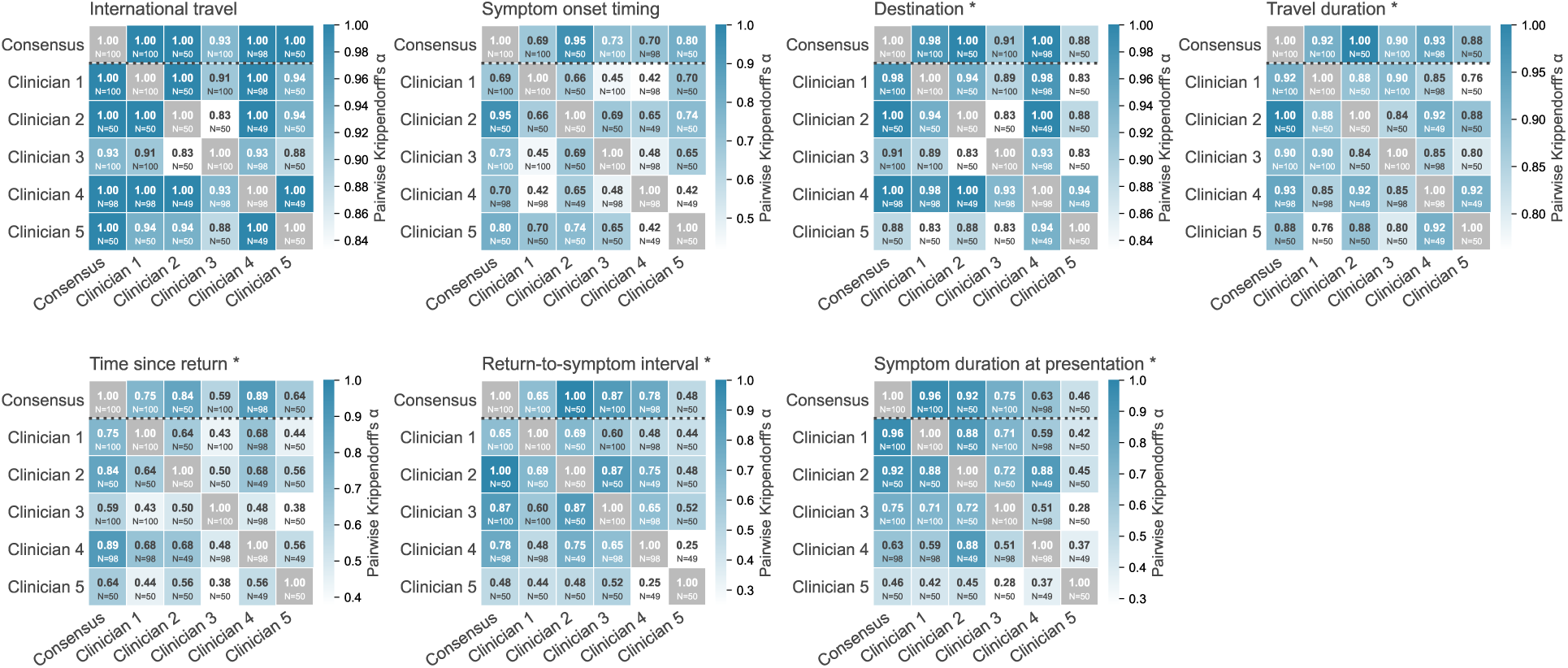
Pairwise agreement among all clinicians. Pairwise Krippendorff’s *α* heatmaps for all seven travel history variables, extending the two examples shown in Figure 2D. Each heatmap shows agreement between the majority-vote consensus and each individual clinician, as well as all pairwise clinician-clinician combinations. Cell counts vary depending on the number of notes each clinician pair annotated in common.

### A.7 Qualitative analysis of clinician disagreement

To further characterise the difference in clinician agreement between travel duration and time since return, we examined presence disagreement for both variables among the same 68 notes for which a clinician majority indicated international travel. Under the annotation guidelines, clinicians were instructed to annotate temporal spans only when explicitly stated in the source text, and not to infer or calculate a duration from other temporal information. Presence disagreement was substantially more frequent for time since return than for travel duration (38/68 [55.9%] versus 10/68 [14.7%]).

Representative examples illustrate the different sources of disagreement (Table 3). For travel duration (row 1-2), disagreement generally involved omission of an explicitly stated duration or differences in the interpretation of a short or imprecise expression. Similar omissions were observed for time since return (row 3-6) even when the relevant interval was stated explicitly. Other cases contained several related temporal statements but did not explicitly state the target interval. Since clinicians were instructed not to infer or calculate durations, these cases were not necessarily annotatable.

**Table 3:** Representative examples of clinician disagreement for travel duration (row 1-2) and time since return (row 3-6). Six notes illustrating different sources of presence and span-boundary disagreement. Excerpts are reproduced from the source reports. **Bold** and <u>underlined</u> text indicate alternative clinician span selections. “Marked by” gives the number of clinicians who annotated a span among those w<u>ho reviewed the note.</u>

| ID | Excerpt | Marked by | Source of disagreement |
| --- | --- | --- | --- |
| 1 | Three weeks before admission, he had traveled to Ethiopia where he had remained for **10 days** without prophylactic chemotherapy against malaria. | C1, C3 (2/3) | Plain omission of an explicit numeral span by 1 of 3 annotators. |
| 2 | Fever started 12 days after a **short** <u>family stay</u> in Northern Italy. | C1, C3 (2/5) | Vague relative phrase with no explicit unit; annotators disagreed on boundary, 3/5 skipped it entirely. |
| 3 | Symptoms began 8 days before his return to the United States. **Two days** after his return, he was admitted to the hospital with hypotension. | C1, C2 (2/5) | Clean, explicit, self-contained sentence, yet only 2/5 marked it – disagreement here is not purely a function of textual explicitness. |
| 4 | He returned to Korea from Germany. On the **tenth day** after the travel, he <u>experienced fever/general ache, severe anal pain, and generalized skin rash every day, and reported to the public health authorities for a voluntary mpox test on the fourth day of symptom onset.</u> | C1, C5 (2/5) | Ambiguous temporal cue embedded within a symptom-focused sentence; 3/5 did not mark it, 1 over-selected the whole clause. |
| 5 | **The pain initially started four days after an elective laparoscopic sleeve gastrectomy performed in Tijuana, Mexico. [...] Ten days later, she visited her primary care provider [...] She returned to the emergency room within one week of her primary care visit.** | C5 (1/5) | No explicit “time since return” expression is stated. The relevant timing is distributed across several sequential relative-time statements; 4/5 annotators did not mark a span, while one selected the wider temporal sequence. |
| 6 | <u>Referred to our hospital from another in May 2013 to investigate a probable leukodystrophy. She was a political refugee from the Democratic Republic of Congo. Before arriving in France in 2010, she had spent 3 years in Angola (Bengo).</u> | C4, C5 (2/5) | No clear “time since return” span, with additional ambiguity over whether the refugee’s movement to France represents international travel; annotators selected different temporal information. |

### A.8 Precision-recall trade off across all variables

Extends the two representative precision-recall plots in Figure 3C to all seven variables (Figure 11), showing per-model precision/recall against the clinician-majority reference across the full variable set.

**Figure 11:**
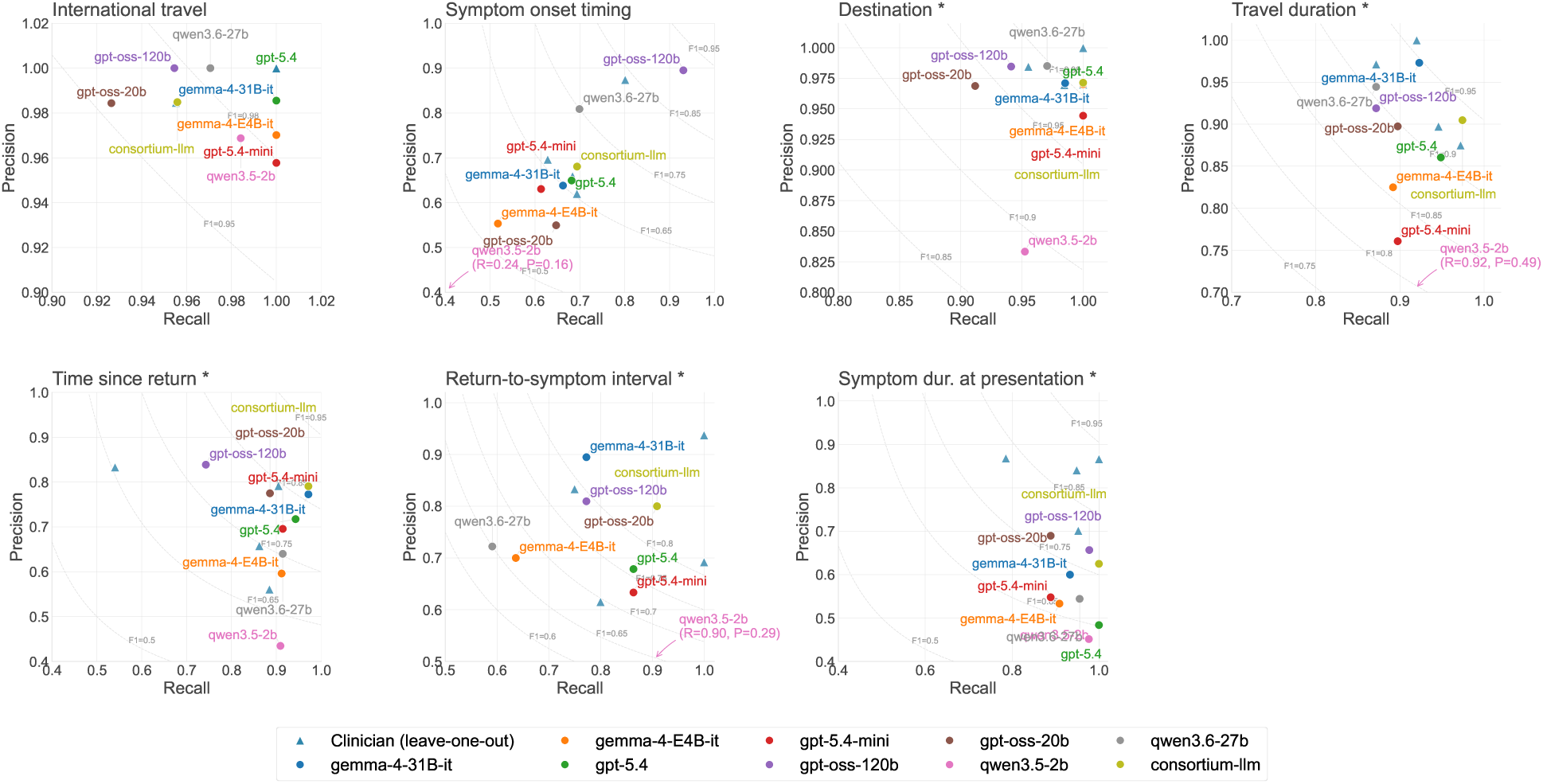
Precision–recall trade-off across all variables. Precision-recall plots for all seven variables, extending the two examples shown in Figure 3C. Each point shows one LLM’s precision and recall against the clinician-majority reference; triangles show individual clinician leave-one-out values. Iso-F1 contours are overlaid for reference.

### A.9 LLM span comparison (strict) against clinicians

Repeats the relaxed span-boundary comparison of Figure 3D under a stricter, exact-match criterion (Figure 12), showing that absolute performance drops substantially for all models and variables once boundary tolerance is removed, though the relative ordering of variables and models is largely preserved.

**Figure 12:**
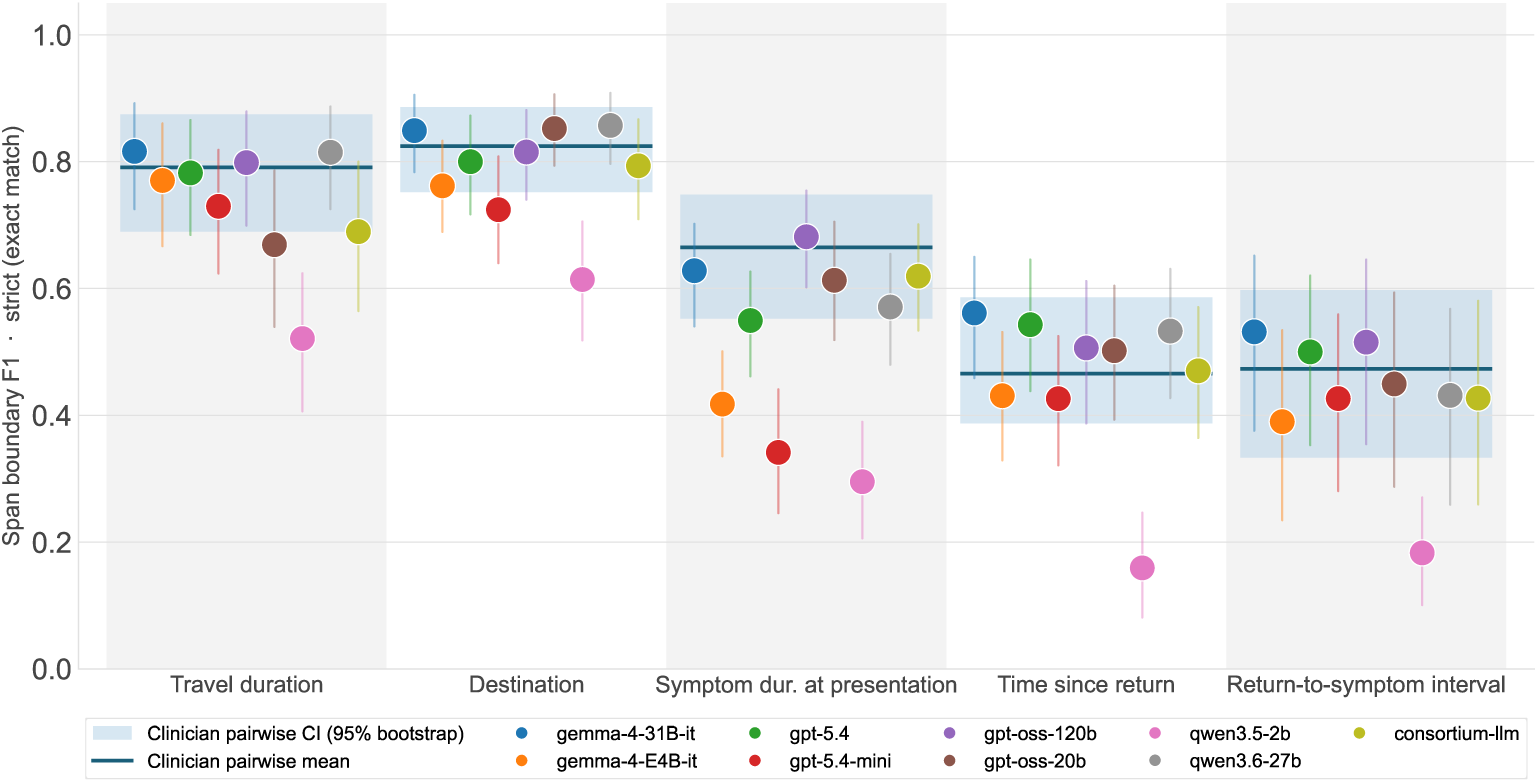
LLM span comparison (strict) against clinicians. Span boundary F1 for all five span variables under strict (exact character-offset match) criteria, contrasting with the relaxed matching (Jaccard *≥* 0.5) reported in Figure 3D. Solid line and shaded band show the mean and 95% bootstrap CI of pairwise clinician span overlap under the same strict criterion.

